# Antibody responses to a suite of novel serological markers for malaria surveillance demonstrate strong correlation with clinical and parasitological infection across seasons and transmission settings in The Gambia

**DOI:** 10.1101/2020.07.10.20067488

**Authors:** Lindsey Wu, Julia Mwesigwa, Muna Affara, Mamadou Bah, Simon Correa, Tom Hall, James G. Beeson, Kevin K.A. Tetteh, Immo Kleinschmidt, Umberto D’Alessandro, Chris Drakeley

## Abstract

**Background:** As malaria transmission declines, sensitive diagnostics are needed to evaluate interventions and monitor transmission. Serological assays measuring malaria antibody responses offer a cost-effective detection method to supplement existing surveillance tools.

**Methods:** A prospective cohort study was conducted from 2013 to 2015 in 12 villages across five administrative regions in The Gambia. Serological analysis included samples from the West Coast Region at the start and end of the season (July and December 2013) and from the Upper River Region in July and December 2013, and April and December 2014. Antigen-specific antibody responses to eight *Plasmodium falciparum* (*P.falciparum*) antigens – Etramp5.Ag1, GEXP18, HSP40.Ag1, Rh2.2030, EBA175 RIII-V, *Pf*MSP1_19_, *Pf*AMA1, *Pf*GLURP.R2 - were quantified using a multiplexed bead-based assay. The association between antibody responses and clinical and parasitological endpoints was estimated at the individual, household, and population level.

**Results:** Strong associations were observed between clinical malaria and concurrent sero-positivity to Etramp5.Ag1 (aOR 4.60 95%CI 2.98 - 7.12), *Pf*MSP1_19_ (aOR 4.09 95%CI 2.60 - 6.44), *Pf*AMA1 (aOR 2.32 95%CI 1.40 - 3.85) and *Pf*GLURP.R2 (aOR 3.12, 95%CI 2.92 – 4.95), while asymptomatic infection was associated with sero-positivity to all antigens. Village-level sero-prevalence amongst children 2-10 years against Etramp5.Ag1, HSP40.Ag1, and *Pf*MSP1_19_ showed the highest correlations with clinical and *P.falciparum* infection incidence rates. For all antigens, there was increased odds of asymptomatic *P.falciparum* infection in subjects residing in a compound with greater than 50% sero-prevalence, with a 2- to 3-fold increase in odds of infection associated with Etramp5.Ag1, GEXP18, Rh2.2030, *Pf*MSP1_19_ and *Pf*AMA1. For individuals residing in sero-positive compounds, the odds of clinical malaria were reduced, suggesting a protective effect.

**Conclusions:** At low transmission, long-lived antibody responses could indicate foci of malaria transmission that have been ongoing for several seasons or years. In settings where sub-patent infections are prevalent and fluctuate below the detection limit of polymerase chain reaction (PCR), the presence of short-lived antibodies may indicate recent infectivity, particularly in the dry season when clinical cases are rare. Serological responses may reflect a persistent reservoir of infection, warranting community-targeted interventions if individuals are not clinically apparent but have the potential to transmit. Therefore, serological surveillance at both the individual and household level may be used to target interventions where there are foci of asymptomatically infected individuals.

## Background

As malaria transmission declines, the prevalence of infection becomes increasingly heterogeneous and focal. In low transmission settings, highly sensitive diagnostics are needed to measure subtle changes in malaria incidence, evaluate the effectiveness of community-based interventions, and monitor potential re-introduction after elimination. Evidence suggests that infections below the detection limit of microscopy and rapid diagnostic tests (RDTs) contribute to on-going transmission, but the magnitude of their public health importance is yet to be determined.(1–3) Nucleic acid amplification tests (NAATs), such as ultrasensitive quantitative polymerase chain reaction (qPCR), can detect less than 1 parasite per microlitre(4,5), but are still sensitive to fluctuations in parasite density during the course of an infection.

Serological assays measuring antibody responses may be a more stable diagnostic alternative, offering a cost-effective detection method to supplement existing surveillance tools. A number of studies in The Gambia have observed spatiotemporal variations in antibody responses to malaria over time(6,7), with similar observations in Tanzania(8), Equatorial Guinea(9), and South Africa(10). The majority of these studies measure long-lived antibody responses, which can persist for years in the absence of re- infection. By contrast, new serological markers of recent malaria exposure have been developed based on antigens that elicit shorter-lived antibody responses and may be capable of measuring changes in malaria transmission dynamics over periods as short as 1-2 years(11,12). These markers require validation with population-representative data from a range of endemic settings and longitudinal cohort studies provide a unique opportunity for a multi-metric assessment of transmission dynamics using clinical, parasitological, and serological endpoints.

Using a suite of novel serological markers of malaria exposure, this study aimed to determine the correlation of antibody responses with clinical and parasitological endpoints at the individual, household, and population level in rural Gambia over two years, and estimate the strength of association between serological markers and gold standard metrics of active malaria infection. These findings will support the utility of serology in measuring recent malaria transmission and allow selection of markers most suitable for use in future surveillance, particularly for reactive detection strategies in pre- and post-elimination settings.

## Methods

### Data and sampling

To understand the dynamics of malaria infection and the impact of annual mass drug administration (MDA), a prospective cohort study was conducted from 2013 to 2015 in 12 villages across five administrative regions - West Coast (WCR), North Bank (NBR), Lower River (LRR), Central River (CRR) and Upper River (URR) Regions, as described by Mwesigwa et al.(13) *Plasmodium falciparum* (*Pf*) prevalence measured by polymerase chain reaction (PCR) ranged from 2.27 to 19.60% in the Central River and Upper River Regions respectively (Figure 1). Residents more than six months of age were enrolled in the study, and monthly surveys were conducted during malaria transmission season from June to December each year, as well as surveys in the dry season in April 2014, and prior to the implementation of MDA in May and June 2014 and 2015 (Figure 2). Individual finger prick blood samples were collected for haemoglobin estimation and on filter paper (Whatman 3 Corporation, Florham Park, NJ, USA) for molecular and serological analysis. Clinical malaria cases included individuals presenting with symptoms at health facilities (e.g., passive case detection) or individuals identified in villages by study nurses with history of fever in the previous 24 hours or axillary temperature ≥ 37.5°C and a positive Rapid Diagnostic Test (RDT) result (Paracheck *Pf*, Orchid Biomedical System, India).

**Figure 1.**
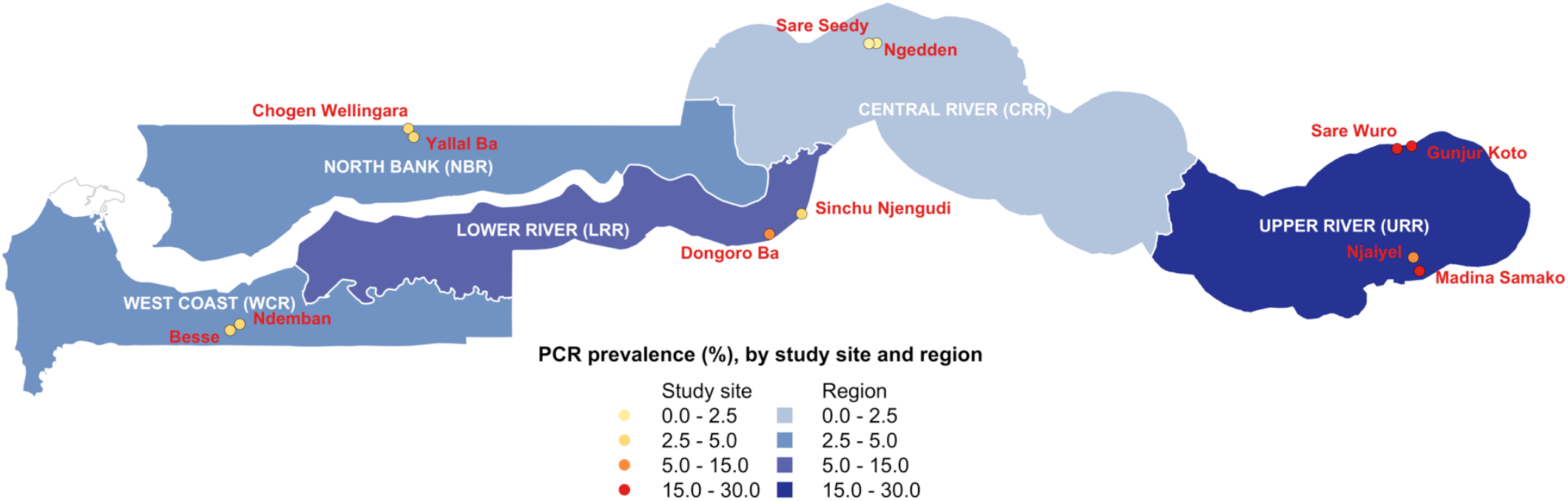
Map of Malaria Transmission Dynamics Study with regions and study villages by PCR prevalence.

**Figure 2.**
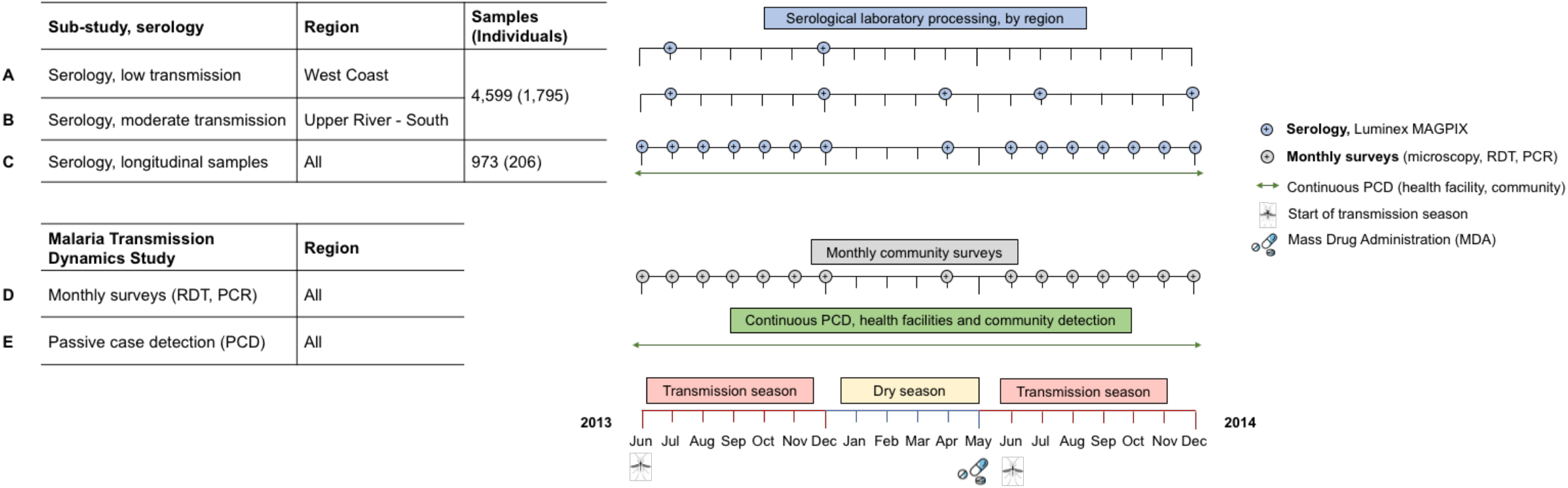
Study timelines. Malaria Transmission Dynamics Study timeline shown in black and green. Serological study timeline shown in blue for West Coast and Upper River Regions (low and moderate transmission settings, respectively). Serological analysis was conducted on samples from whole-village monthly surveys in N’demban and Besse in the West Coast Region (A), Njaiyal and Madina Samako in the Upper River Region (B) and longitudinal samples from individuals with a positive rapid diagnostic test (RDT) or polymerase chain reaction (PCR) test result during the Malaria Transmission Dynamics Study. Samples for serological analysis were processed on the Luminex MAGPIX, samples from monthly surveys were analysed using microscopy, rapid diagnostic tests (RDTs) and polymerase chain reaction (PCR).

The serological study presented here is a subset of the Malaria Transmission Dynamics Study and included all available samples (n=4,599) from a selection of monthly surveys in four villages, totalling 1,795 individuals (Figure 2). In the West Coast Region (Besse and N’Demban), samples processed for serological analysis were from surveys conducted at the start of the transmission season in July 2013 (N=534) and at the end of the season in December 2013 (N=524). In the Upper River Region (URR), serological analysis included all samples collected in Njaiyal and Madino Samako in July 2013 (N=778), December 2013 (N=628), April (dry season) 2014 (N=799), and December 2014 (N=737) (Table 1). These regions represent extremes of two transmission intensities, with months selected at the start and end of the transmission season. Samples from clinical PCD cases were linked by study participant identification code to samples from the same individuals collected during routine monthly surveys. To further estimate the association between individual-level antibody responses and concurrent clinical or *Pf* infection, whole-village monthly survey samples in the West Coast Region and Upper River Region as described above were combined with an additional subset of 973 longitudinal samples from 206 individuals who experienced a positive RDT or PCR test result at any point during the Malaria Transmission Dynamics Study (Figure 2). For these individuals, all available samples from the study were processed to longitudinally capture their serological responses before and after a positive RDT or PCR test result.

**Table 1.** Summary of antigens in multiplex Luminex panel.

| Gene ID | Antigen name | Strain | Antigen bead coupling concentration (ug/mL) | Purification Tag | Location | Description | Reference |
| --- | --- | --- | --- | --- | --- | --- | --- |
| PF3D7_0930300 | *Pf* MSP1.19 | Wellcome | 42.31 | GST | Merozoite surface | 19kDa fragment of MSP1 molecule | (45) |
| PF3D7_1133400 | *Pf* AMA1 | FVO | 3.90 | His<sub>x6</sub> | Sporozoite / Merozoite | Apical membrane antigen 1 | (46) |
| PF3D7_1035300 | *Pf* GLURP.R2 | F32 | 9.22 | N/A | Merozoite | Glutamate rich protein R2 | (47) |
| PF3D7_0731500 | EBA175 RIII-V | 3D7 | 408.32 | GST | Merozoite | Erythrocyte binding antigen-175 | (48) |
| PF3D7_1335400 | Rh2.2030 | D10 | 244.30 | GST | Merozoite | Reticulocyte binding protein homologue 2 | (49) |
| PF3D7_0532100 | Etramp5.Ag1 | 3D7 | 34.93 | GST | iRBC/PVM | Early transcribed membrane protein 5 | (50) |
| PF3D7_0402400 | GEXP18 | 3D7 | 625.00 | GST | Gametocytes | Gametocyte exported protein 18 | (11) |
| PF3D7_0501100.1 | HSP40.Ag1 | 3D7 | 42.54 | GST | iRBC / Gametocytes | Heat shock protein 40, type II | (11) |
| -- | GST | -- | 85.99 | -- | -- | GST expression tag |  |
| -- | TT | -- | 61.52 | -- | -- | Tetanus Toxoid |  |

\*infected red blood cell – iRBC; parasitophorous vacuole membrane – PVM, glutathione S –transferase - GS

This study was approved by the Gambia Government/MRC Joint Ethics Committee (SCC1318). Verbal consent was first obtained at village sensitisation meetings, followed by individual written informed consent for all participants. Parents/guardians provided written consent for children less than 17 years, and assent was obtained from children between 12 and 17 years.

### Antigen selection and design

Selection of antigens was based on an initial screen of 856 candidates on an *in vitro* transcription and translation (IVTT) protein microarray based on correction with malaria infection in children.(11) Antigens were generated and expressed in *Escherichia coli* (*E.coli*) as glutathione S-transferase (GST)- tagged fusion proteins(14–16), with the exception of *Pf*AMA1 expressed in Pichia pastoris as a histidine-tagged protein.(17) Protein purification was conducted by affinity chromatography (Glutathione Sepharose 4B, GE Healthcare Life Sciences) or HisPur Ni-NTA (Invitrogen) for GST- and His-tagged proteins, respectively, and concentration, quality, and purity of antigen yield assessed using a Bradford assay and SDS-PAGE. Bacterial lysate from culture of untransformed *E.coli* was used in assay buffers to eliminate background reactivity *E.coli* proteins that is not specific to malaria target proteins.

Additionally, to account for potential non-malaria reactivity against GST-tagged fusion proteins, GST-coupled beads were included to quantify GST-specific immunoglobulin (IgG) responses and correct for potential non-malaria specific binding. After laboratory processing, there were 71 participant samples with GST antibody responses above 1,000 MFI, which was defined as the threshold to indicate potential non-malaria specific binding. These 71 samples were excluded from further analyses to avoid the influence of non-malaria specific responses. Tetanus toxoid (TT, Massachusetts Biologic Laboratories) was also included as an internal positive control, assuming that vaccinated Gambians would show antibody responses to this protein target. A summary of antigen constructs and coupling conditions are detailed in Table 1.

### Laboratory procedures

Antigen-specific antibody responses were quantified using the Luminex MAGPIX protocol described in Wu et al 2019.(18) Plasma was eluted from 6mm dried blood spots (DBS) (4 μl whole blood equivalent) and shaken overnight at room temperature in 200 μl of protein elution buffer containing phosphate buffered saline (PBS) (pH 7.2), 0.05% sodium azide and 0.05% Tween-20, yielding an initial 1:50 sample dilution. One day prior to assay processing, samples were diluted to a final 1:500 dilution using 10 μl of the 1:50 pre-dilution sample and 90 μl of blocking Buffer B to prevent non-specific binding (1xPBS, 0.05% Tween, 0.5% bovine serum albumin (BSA), 0.02% sodium azide, 0.1% casein, 0.5% polyvinyl alcohol (PVA), 0.5% polyvinyl pyrrolidone (PVP) and 1,500 μg/ml *E.coli* extract). Negative and positive controls were also incubated one day prior in Buffer B, with negative controls prepared at a 1:500 dilution and Gambian pooled positive controls in a 6-point 5-fold serial dilution (1:10-1:31,250). The positive control was based on a pool of 22 serum samples from malaria hyper-immune individuals in The Gambia, and ten individual plasma samples from European malaria-naive adults were used as negative controls.

Samples were prepared for diagnostic PCR as described by Mwesigwa et al 2017.(13) Briefly, DNA was extracted from three 6-mm DBS using the automated QIAxtractor robot (Qiagen). Negative and positive (3D7) controls were included to control for cross contamination and DNA extraction efficiency, respectively. The DBS were lysed by incubation in tissue digest buffer at 60 °C for 1 hour and digested eluates were applied onto capture plates, washed, and the DNA eluted into 80 µl. The extracted DNA (4 µl) was used in a nested PCR, amplifying the multi-copy *Plasmodium* ribosomal RNA gene sequences using genus and species specific primers.(19) All PCR products were run using the QIAxcel capillary electrophoresis system (Qiagen), using the screening cartridge and 15-1,000 bp- alignment marker. Results were exported and double scored using both the QIAxcel binary scoring function and manually by visualization of the gel images and discrepancies were scored by a third independent reader. All readers were blinded to participant survey data.

### Statistical analyses

Data analysis was based on total immunoglobulin (IgG) levels to five antigens as potential markers of sero-incidence(11) - early transcribed membrane protein 5 (Etramp5.Ag1), gametocyte export protein 18 (GEXP18), heat shock protein 40 (HSP40.Ag1), erythrocyte binding antigen 175 RIII-V (EBA175), reticulocyte binding protein homologue 2 (Rh2.2030). Three antigens associated with long-lived antibody response – *P.falciparum* merozoite surface antigen 1 19-kDa carboxy-terminal region (*Pf*MSP1_19_), *P.falciparum* apical membrane antigen 1 (*Pf*AMA1), and *P.falciparum* glutamate-rich protein, region 2 (*Pf*GLURP.R2) – were included as a comparison, both historically used to assess sero- conversion rates over time(8,9) and previously studied in The Gambia(6,7). For antigens associated with long-lived antibodies (*Pf*MSP1_19_, *Pf*AMA1, *Pf*GLURP.R2), individuals residing in an endemic region previously exposed to malaria but not recently infected may still have residual antibody levels that are significantly higher than malaria-naïve individuals in non-endemic regions. Therefore, a two-component Gaussian mixture model was used to define distributions of negative and positive antibody levels, expressed in units of median fluorescence intensity (MFI). Sero-positivity thresholds were defined as the mean log MFI values plus two standards deviations of the negative distribution.(20) Mixture models were estimated using the ‘*normalmixEM’* function in the *‘mixtools’* package v1.0.4 in R version 3.6.1. For antigens associated with shorter-lived antibodies where statistical evidence of a bimodal distribution of antibody responses in the population was not strong given the more rapid decay of antibody levels post-infection - Etramp5.Ag1, GEXP18, HSP40.Ag1, EBA175, Rh2.2030 - the sero-positivity threshold was defined by the mean log MFI plus three standard deviations of 71 malaria-naïve European blood donors used as negative controls.

Individual-level association between antibody response and the concurrent odds of clinical malaria (passively detected via the health facility or study nurses in the community) or asymptomatic *P.falciparum* infection (actively detected using PCR from monthly survey samples) were assessed using generalised estimating equations (GEE). Analysis was adjusted for age group (1-5 years, 6-15 years, and greater than 15 years) and use of long-lasting insecticide treated nets (LLINs) in the last 24 hours and allowed for clustering at the compound level, where compound is a collection of households centrally located around a main residence. The magnitude of the association between antibody response and odds of infection was evaluated for interaction with age group.

Using a GEE model, individual-level odds of clinical malaria or asymptomatic infection was assessed for association with residence in the same compound as a sero-positive individual. Similarly, the association between individual odds of infection and compound-level sero-prevalence (<50% or >50%), was assessed using a mixed-effects generalised linear model adjusted for age group and LLIN use, with random-effects at the compound-level which also accounts for the potential effect of transmission intensity. To ensure that estimates are not biased by the sero-prevalence of small compounds, analysis was weighted by the number of individuals in the compound (whose sero-status was assessed) and only included compounds with at least four individuals. Models were fit using the ‘geeM’ and ‘lme4’ packages in R version 3.14.

Village-level sero-prevalence amongst children aged 2-10 years in the West Coast Region and Upper River Region in July and December 2013 (n=1,001) was compared against all-age clinical and *P.falciparum* infection incidence rates from the same months, the latter of which were previously reported by Mwesigwa et al(13). Monthly clinical and *P.falciparum* infection incidence rates were defined respectively as the number of new clinical cases or *P.falciparum* infections (PCR-positive individuals who were PCR-negative in the previous monthly survey) divided by total person-years at risk. This age range was selected to align with other routine surveillance metrics such as annual parasite index (API) commonly using this age group as a sentinel population. The strength of the relationship between village-level incidence rates and sero-prevalence for each antigen was assessed using Pearson’s correlation coefficient. Data from April and December 2014 were not included in the analysis because clinical incidence and *Pf* infection rates from these surveys have not yet been reported.

## Results

### Village-level seroprevalence

Data from 2,001 individuals were available for analysis of antibody responses from 5,572 samples between June 2013 and December 2014 (Figure 1). Based on whole-village monthly survey data in the West Coast and Upper River Regions, slightly more than half of participants were female in both the West Coast (55.0%) and the Upper River (54.3%) Regions, and higher LLIN use in the West Coast (96.8%) compared to the Upper River Region (46.6%). In the West Coast Region, *P. falciparum* infection rate in December 2013 (0.67 95%CI 0.40 - 1.13) was three times higher than at the start of the transmission season in July 2013 (0.23 95%CI 0.13 - 0.39). In the Upper River Region, *P. falciparum* infection rate in December 2013 (2.87 95%CI 2.36 - 3.50) was five times higher than July 2013 (0.56 95%CI 0.42 - 0.74) (Table 3). Sero-prevalence amongst 2-10 years olds in the West Coast Region ranged from 1.0% (95%CI 0.0 - 2.3) for Rh2.2030 in July 2013 (start of the transmission season) and 13.7% (95%CI 8.9 - 18.5) for Etramp5.Ag1 in December 2013 (end of transmission season). In the Upper River Region South, sero-prevalence across all antigens was higher, ranging from 5.1% (95%CI 2.5 - 7.7) for *Pf*MSP1_19_ in July 2013 to 36.3% (95%CI 30.2 - 42.3) for GEXP18 in December (Table 3). Sero-prevalence in this age group was highly correlated with both clinical malaria and *P.falciparum* infection incidence rates (Figure 2, Table 4). In the Upper River Region in 2014, no statistically significant differences in sero-prevalence amongst 2-10 year olds were observed for any antigens (Additional File 1 – Table S1). It should be noted that MDA was implemented in May – June 2014, so results may not be comparable to 2013 prior to intervention.

**Table 2.**
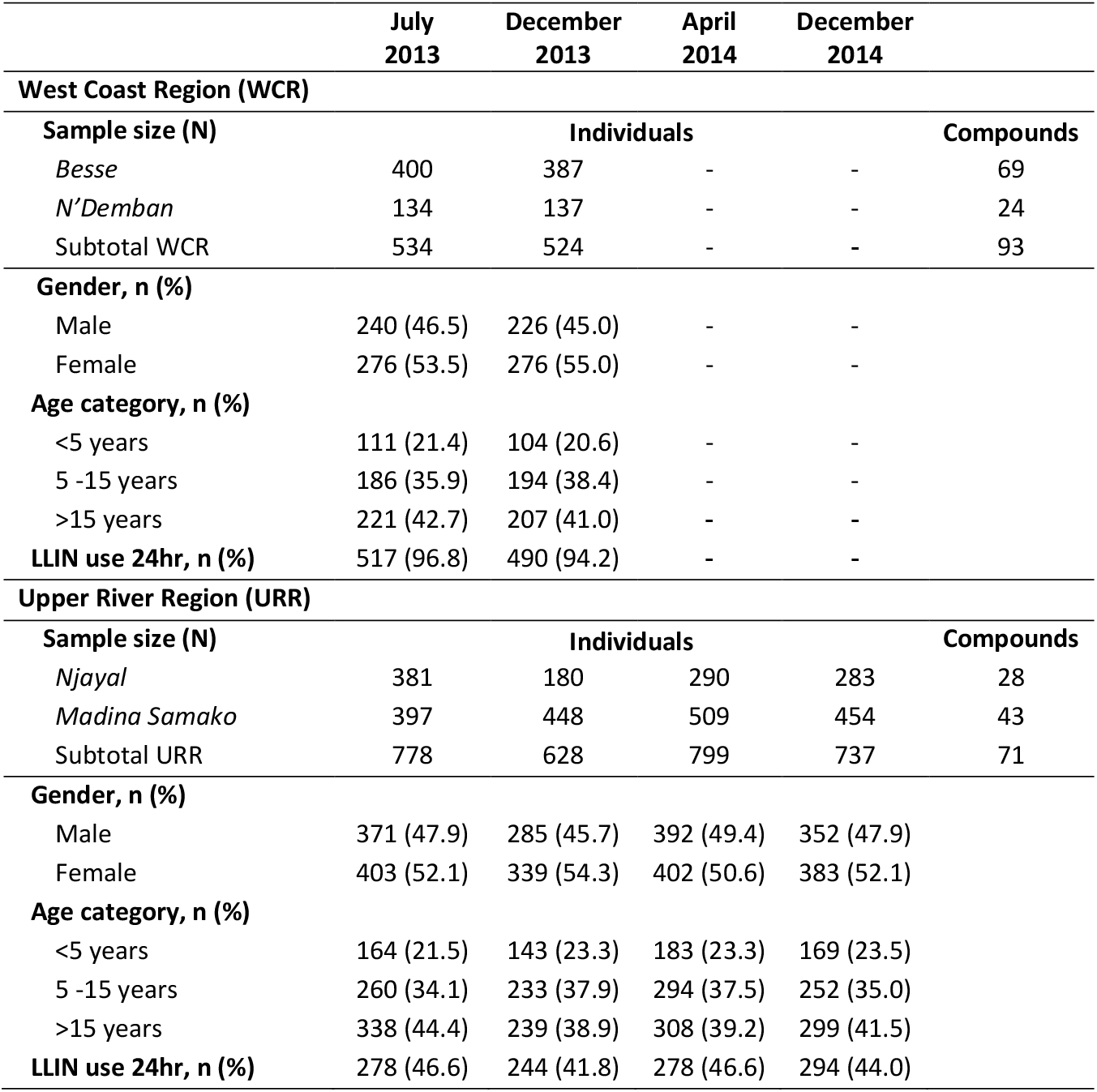
Sample size and study subject characteristics by region and survey month. Sample size (N) reported as number of individuals in each village for each monthly survey and total number of compounds (with compound defined as a collection of households centrally located around a main residence). Study subject characteristics reported as number and percentage individuals of out of total individuals in each monthly survey. Dashed lines indicate survey months where serological analysis was not conducted in the West Coast Region.

**Table 3.** Regional sero-prevalence (ages 2-10 years) by antigen and all-age incidence rates for clinical malaria and *Pf* infection. Mean sero-prevalence (%) and incidence rates with 95%CI shown in parentheses.

|  | West Coast Region (WCR) |  | Upper River Region (URR) |  |
| --- | --- | --- | --- | --- |
|  | July 2013 | December 2013 | July 2013 | December 2013 |
| **Clinical incidence rate** | 0.08 (0.04 - 0.21) | 0.14 (0.04 - 0.44) | 0.13 (0.04 - 0.39) | 0.21 (0.1 - 0.51) |
| ***Pf* infection rate** | 0.23 (0.13 - 0.39) | 0.67 (0.40 - 1.13) | 0.56 (0.42 - 0.74) | 2.87 (2.36 - 3.50) |
| **Sero-prevalence (ages 2-10 years)** |  |  |  |  |
|  | n=205 | n=197 | n=275 | n=324 |
| **Etramp5.Ag1** | 4.4% (1.6 - 7.2) | 13.7% (8.9 - 18.5) | 8.7% (5.4 - 12.1) | 33.3% (27.4 - 39.3) |
| **GEXP18** | 11.7% (7.3 - 16.1) | 12.7% (8.0 - 17.3) | 25.1% (20.0 - 30.2) | 36.3% (30.2 - 42.3) |
| **HSP40.Ag1** | 5.4% (2.3 - 8.5) | 6.1% (2.8 - 9.4) | 8.4% (5.1 - 11.6) | 23.8% (18.4 - 29.1) |
| **Rh2.2030** | 1.0% (0.0 - 2.3) | 1.0% (0.0 - 2.4) | 9.8% (6.3 - 13.3) | 25.4% (19.9 - 30.9) |
| **EBA175** | 1.5% (0.0 - 3.1) | 1.0% (0.0 - 2.4) | 7.3% (4.2 - 10.3) | 15.8% (11.2 - 20.5) |
| ***Pf*MSP1<sub>19</sub>** | 2.9% (0.6 - 5.2) | 8.1% (4.3 - 11.9) | 5.1% (2.5 - 7.7) | 22.1% (16.8 - 27.3) |
| ***Pf*AMA1** | 4.4% (1.6 - 7.2) | 4.1% (1.3 - 6.8) | 17.8% (13.3 - 22.3) | 33.3% (27.4 - 39.3) |
| ***Pf*GLURP.R2** | 4.4% (1.6 - 7.2) | 8.1% (4.3 - 11.9) | 16.4% (12.0 - 20.7) | 30.4% (24.6 - 36.2) |

**Table 4.** Pearson’s correlation coefficients between regional sero-prevalence (ages 2-10 years) and all-age incidence rates for clinical malaria and *Pf* infection. Correlation estimates are based on data from the West Coast Region (WCR) and Upper River Region (URR) in July and December 2013. Correlations excluding data from URR December 2013 are shown in brackets.

| Antigen | Pearson's correlation coefficient |  |
| --- | --- | --- |
|  | Sero-prevalence vs. Clinical incidence rate | Sero-prevalence vs. *Pf* infection incidence rate |
| **Etramp5.Ag1** | 0.97 [0.92] | 0.99 [0.95] |
| **GEXP18** | 0.85 [0.42] | 0.87 [0.34] |
| **HSP40.Ag1** | 0.90 [0.56] | 0.99 [0.48] |
| **Rh2.2030** | 0.88 [0.36] | 0.94 [0.28] |
| **EBA175** | 0.85 [0.29] | 0.91 [0.21] |
| ***Pf*MSP1<sub>19</sub>** | 0.95 [0.90] | 0.99 [0.93] |
| ***Pf*AMA1** | 0.85 [0.34] | 0.89 [0.26] |
| ***Pf*GLURP.R2** | 0.92 [0.62] | 0.93 [0.55] |

In both regions in July and December 2013, correlations between *P.falciparum* infection rate and sero- prevalence were strongest for Etramp5.Ag1 (ρ=0.99), HSP40.Ag1 (ρ=0.99), *Pf*MSP1_19_ (ρ=0.99). and Rh2.2030 (ρ=0.94). Incidence of clinical malaria had the strongest correlations with Etramp5.Ag1 (ρ=0.97), *Pf*MSP1_19_ (ρ=0.95), *Pf*GLURP.R2 (ρ=0.92), and HSP40.Ag1 (ρ=0.90), Correlations with both clinical incidence and *P.falciparum* infection rates remained high for Etramp5.Ag1 (ρ=0.92 and ρ=0.95, respectively) and *Pf*MSP1_19_ (ρ=0.90 and ρ=0.93, respectively), after excluding data from December 2013 from the Upper River Region South as a potential outlier. These correlations are based on few datapoints, with three observations when excluding the Upper River Region in December 2013.

### Individual-level association between clinical malaria, asymptomatic *P.falciparum* infection and concurrent sero-positivity

The odds of clinical malaria were higher in individuals sero-positive to six (out of eight) of the antigens analysed (Etramp5.Ag1, GEXP18, Rh2.2030, *Pf*MSP1_19_, *Pf*AMA1, and *Pf*GLURP.R2) (Figure 3A, Table 5). After adjusting for age and LLIN use, clinical malaria was associated with concurrent sero-positivity to three antigens (Etramp5.Ag1, *Pf*MSP1_19,_ *Pf*AMA1, and *Pf*GLURP.R2), while odds of asymptomatic infection were strongly associated with sero-positivity to all antigens. Amongst individuals sero- positive for Etramp5.Ag1, there was increased odds of both clinical malaria (aOR 4.60, 95%CI 2.98 - 7.12, p<0.001) and asymptomatic infection (aOR 3.33, 95%CI 2.72 - 4.08, p<0.001) (Table 5). Association with clinical malaria was slightly lower in individuals sero-positive for *Pf*MSP1_19_ (aOR 4.09, 95%CI 2.60 - 6.44, p<0.001), *Pf*GLURP.R2 (aOR 3.12, 95%CI 2.12 - 4.59, p<0.001) and *Pf*AMA1 (aOR 2.32, 95%CI 1.40 - 3.845 p=0.001). Odds of asymptomatic infection in individuals sero-positive to GEXP18 (aOR 3.12, 95%CI 2.50 – 3.90, p<0.001), Rh2.2030 (aOR 3.06, 95%CI 2.40 - 3.89, p<0.001), *Pf*AMA1 (aOR 3.80, 95%CI 2.95 - 4.90, p<0.001), and *Pf*GLURP.R2 (aOR 3.80 95%CI 2.91 – 4.95, p<0.001) was similar to Etramp5.Ag1.

**Table 5.** Odds of clinical malaria and asymptomatic infection by individual serological status, unadjusted and adjusted for age group (<5 years, 5-15 years, and > 15 years) and LLIN use in the last 24 hours.

| **Odds of clinical malaria or asymptomatic infection amongst sero-positive individuals, by antigen** |  |  |  |  |  |  |
| --- | --- | --- | --- | --- | --- | --- |
|  | **OR** | **Unadjusted**<br>**95%CI** | **p-value** | **aOR** | **Adjusted**<br>**95%CI** | **p-value** |
| **Etramp5.Ag1** |  |  |  |  |  |  |
| *Clinical malaria* | 5.88 | 3.44 - 10.03 | <0.001 | 4.60 | 2.98 - 7.12 | <0.001 |
| *Asymptomatic infection* | 3.20 | 2.64 - 3.88 | <0.001 | 3.33 | 2.72 - 4.08 | <0.001 |
| **GEXP18** |  |  |  |  |  |  |
| *Clinical malaria* | 2.57 | 1.44 - 4.61 | 0.002 | 1.48 | 0.90 - 2.41 | 0.122 |
| *Asymptomatic infection* | 2.94 | 2.38 - 3.64 | <0.001 | 3.12 | 2.50 - 3.90 | <0.001 |
| **HSP40.Ag1** |  |  |  |  |  |  |
| *Clinical malaria* | 1.67 | 0.88 - 3.18 | 0.119 | 0.99 | 0.59 - 1.65 | 0.956 |
| *Asymptomatic infection* | 2.53 | 2.06 - 3.10 | <0.001 | 2.64 | 2.09 - 3.33 | <0.001 |
| **Rh2.2030** |  |  |  |  |  |  |
| *Clinical malaria* | 2.20 | 1.10 - 4.38 | 0.025 | 1.28 | 0.77 - 2.12 | 0.338 |
| *Asymptomatic infection* | 2.45 | 1.98 - 3.03 | <0.001 | 3.06 | 2.40 - 3.89 | <0.001 |
| **EBA175** |  |  |  |  |  |  |
| *Clinical malaria* | 1.37 | 0.67 - 2.81 | 0.390 | 1.54 | 0.86 - 2.75 | 0.147 |
| *Asymptomatic infection* | 2.09 | 1.74 - 2.51 | <0.001 | 2.86 | 2.21 - 3.72 | <0.001 |
| **PfMSP1<sub>19</sub>** |  |  |  |  |  |  |
| *Clinical malaria* | 3.83 | 1.95 - 7.51 | <0.001 | 4.09 | 2.60 - 6.44 | <0.001 |
| *Asymptomatic infection* | 2.29 | 1.82 - 2.88 | <0.001 | 2.49 | 1.90 - 3.27 | <0.001 |
| **PfAMA1** |  |  |  |  |  |  |
| *Clinical malaria* | 2.21 | 1.25 - 3.90 | 0.006 | 2.32 | 1.40 - 3.85 | 0.001 |
| *Asymptomatic infection* | 2.62 | 2.12 - 3.24 | <0.001 | 3.80 | 2.95 - 4.90 | <0.001 |
| **PfGLURP.R2** |  |  |  |  |  |  |
| *Clinical malaria* | 2.17 | 1.28 - 3.68 | 0.004 | 3.12 | 2.12 - 4.59 | <0.001 |
| *Asymptomatic infection* | 2.38 | 1.96 - 2.89 | <0.001 | 3.80 | 2.92 - 4.95 | <0.001 |

**Figure 3.**
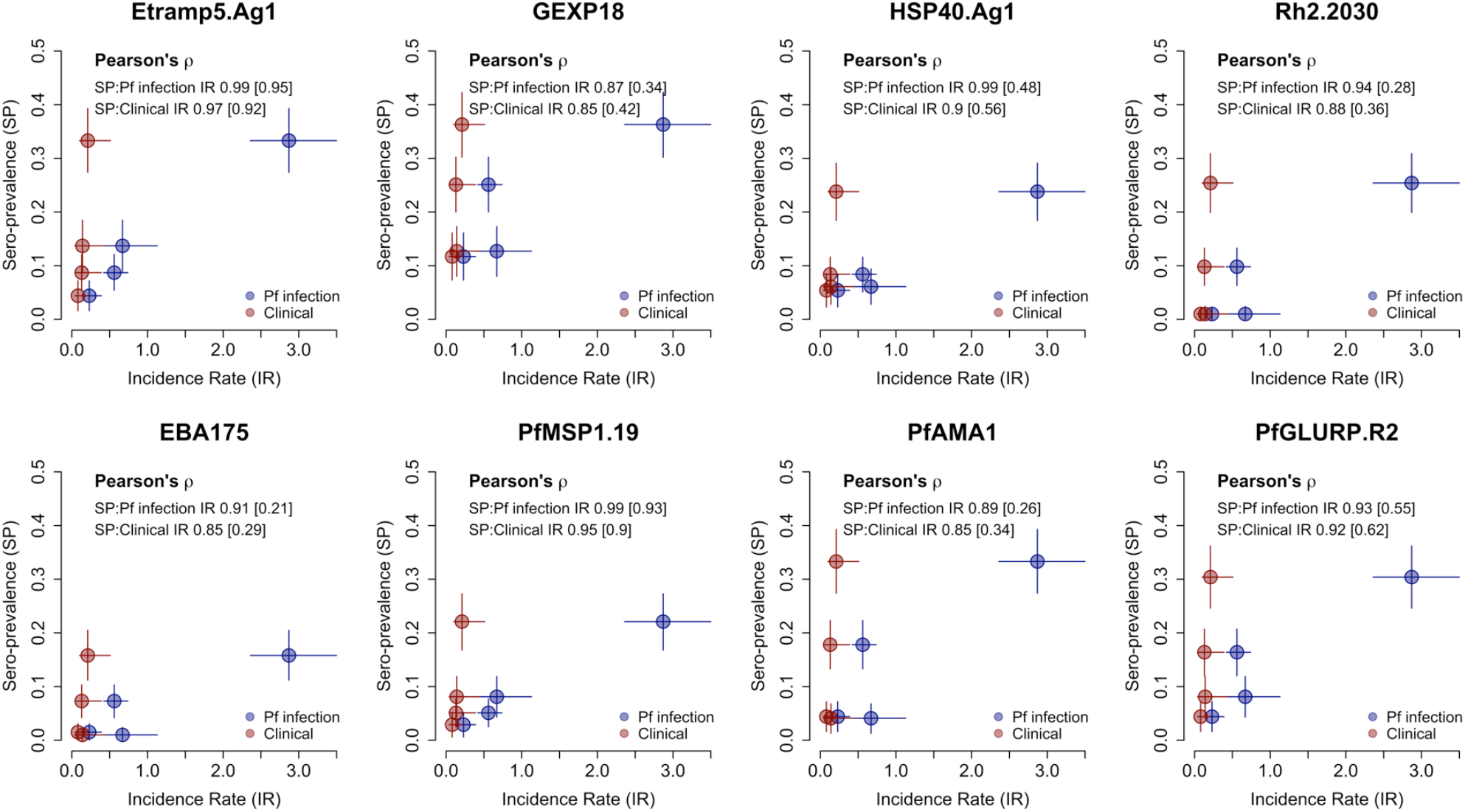
Pearson’s correlation between regional sero-prevalence (ages 2-10 years) and all-age incidence rates for clinical malaria and Pf infection. Correlations between sero-prevalence and incidence rates for clinical malaria (new clinical episodes divided by person-years at risk) in red and Pf infection (new PCR infections divided by person- years at risk) In blue were based on data from the West Coast Region (WCR) and the Upper River Region (URR) in July and December 2013. Correlations excluding data from URR December 2013 shown in brackets.

### Individual-level odds of clinical malaria, asymptomatic infection when residing in compounds with sero-positive individuals

The odds of clinical malaria were reduced for individuals residing in the same compound with an individual sero-positive to any of the seven antigens (Figure 3B, Table 6), from an 83% reduction if residing in the same compound as an individual sero-positive for Etramp5.Ag1 (aOR 0.17, 95%CI 0.03- 0.84, p=0.036) to a 95% reduction if residing in the same compound as someone sero-positive for *Pf*AMA1 (aOR 0.05, 95%CI 0.01 - 0.52, p=0.011). Conversely, the odds of asymptomatic infection were positively associated with residing in the same compound as sero-positive individuals for five antigens (Etramp5.Ag1, GEXP18, Rh2.2030, *Pf*MSP1_19,_ and *Pf*AMA1). Adjusted odds of asymptomatic infection was nearly 3-fold higher if residing with an individual sero-positive for Etramp5.Ag1 (aOR 2.87, 95%CI 1.62 - 5.07, p<0.001) and for GEXP18 (aOR 2.61, 95%CI 1.54 - 4.42, p<0.001). If residing in a compound with individuals sero-positive to Rh2.2030, *Pf*MSP1_19_ or *Pf*AMA1, the odds of asymptomatic infection were all nearly twice as high as individuals living in compounds sero-negative to these antigens (Table 6).

**Table 6.** Odds of clinical malaria and asymptomatic *Pf* infection by compound serological status. Estimates are weighted by compound size and shown as unadjusted odds ratios (OR) and adjusted odds ratios (aOR) for age group (<5 years, 5-15 years, and > 15 years).

| **Odds of clinical malaria or asymptomatic *Pf* infection for individuals residing in sero-positive compounds** |  |  |  |  |  |  |
| --- | --- | --- | --- | --- | --- | --- |
|  | **OR** | **Unadjusted**<br>**95%CI** | **p-value** | **aOR** | **Adjusted**<br>**95%CI** | **p-value** |
| **Etramp5.Ag1** |  |  |  |  |  |  |
| *Clinical malaria* | 0.19 | 0.03 - 1.06 | 0.059 | 0.17 | 0.03 - 0.89 | 0.036 |
| *Asymptomatic infection* | 2.83 | 1.62 - 4.96 | <0.001 | 2.87 | 1.62 - 5.07 | <0.001 |
| **GEXP18** |  |  |  |  |  |  |
| *Clinical malaria* | 0.13 | 0.02 - 0.90 | 0.039 | 0.11 | 0.02 - 0.73 | 0.022 |
| *Asymptomatic infection* | 2.65 | 1.58 - 4.47 | <0.001 | 2.61 | 1.54 - 4.42 | <0.001 |
| **HSP40.Ag1** |  |  |  |  |  |  |
| *Clinical malaria* | 0.14 | 0.02 - 0.80 | 0.027 | 0.13 | 0.02 - 0.70 | 0.018 |
| *Asymptomatic infection* | 1.32 | 0.77 - 2.28 | 0.316 | 1.38 | 0.79 - 2.41 | 0.257 |
| **Rh2.2030** |  |  |  |  |  |  |
| *Clinical malaria* | 0.07 | 0.01 - 0.48 | 0.006 | 0.06 | 0.01 - 0.35 | 0.002 |
| *Asymptomatic infection* | 1.69 | 0.96 - 2.99 | 0.070 | 1.92 | 1.10 - 3.36 | 0.022 |
| **EBA175** |  |  |  |  |  |  |
| *Clinical malaria* | 0.07 | 0.01 - 0.61 | 0.016 | 0.07 | 0.01 - 0.57 | 0.013 |
| *Asymptomatic infection* | 1.00 | 0.50 - 2.01 | 0.992 | 1.02 | 0.50 - 2.09 | 0.954 |
| **PfMSP1<sub>19</sub>** |  |  |  |  |  |  |
| *Clinical malaria* | 0.15 | 0.03 - 0.87 | 0.034 | 0.12 | 0.03 - 0.50 | 0.004 |
| *Asymptomatic infection* | 1.87 | 1.15 - 3.06 | 0.012 | 1.95 | 1.19 - 3.20 | 0.008 |
| **PfAMA1** |  |  |  |  |  |  |
| *Clinical malaria* | 0.06 | 0.01 - 0.62 | 0.018 | 0.05 | 0.01 - 0.52 | 0.011 |
| *Asymptomatic infection* | 1.96 | 1.01 - 3.81 | 0.046 | 1.99 | 1.00 - 3.93 | 0.048 |
| **PfGLURP.R2** |  |  |  |  |  |  |
| *Clinical malaria* | 0.06 | 0.01 - 0.53 | 0.012 | 0.05 | 0.01 - 0.43 | 0.006 |
| *Asymptomatic infection* | 1.59 | 0.85 - 2.97 | 0.145 | 1.52 | 0.77 - 3.01 | 0.228 |

The prevalence of sero-positive compounds for long-lived antibody responses varied by antigen and region but remained generally stable throughout the transmission season. For *Pf*MSP1_19_, prevalence of sero-positive compounds was 32.2% (95%CI 14.9 – 49.5) in July 2013 and 39.8% (95%CI 23.6 – 56.0) in December 2013, while prevalence of compounds sero-positive for *Pf*AMA1 and *Pf*GLURP.R2 ranged between 77.0% (95%CI 66.9 – 87.1) in July 2013 to 80.7% (95%CI 71.5 – 89.9) in December 2013 (Additional File 1 – Table S1). In the Upper River Region, nearly all compounds were sero-positive to *Pf*AMA1 and *Pf*GLURP.R2, from 96.2% (95%CI 91.2 – 100) in July 2013 to 100% in December 2014, but prevalence was slightly lower for *Pf*MSP1_19,_ which was between 70.4% (95%CI 55.9 – 84.9) in July 2013 and 88.6% (95%CI 80.7 – 96.5) in December 2014, By contrast, prevalence of sero-positive compounds for short-lived antibody responses were lower in both regions, ranging from 41.4% (95%CI 25.3 – 57.5) for Etramp5.Ag1 and 63.3% (95%CI 50.5 – 76.0) for EBA175 in July 2013, and from 72.2% (95%CI 58.2 − 86.3) for Etramp5.Ag1 in July 2013 to 97.1% (95%CI 93.2 – 100) for GEXP18 in December 2014.

### Association between individual-level clinical malaria, asymptomatic infection, and compound sero- prevalence

When stratifying by compound-level sero-prevalence, mean regression estimates found a reduced odds of clinical malaria if residing in compound with sero-prevalence less than 50% for all antigens, and an increased odds of clinical malaria in compounds with greater than 50% sero-prevalence for four antigens (Etramp5.Ag1, GEXP18, HSP40.Ag1, and *Pf*MSP1_19_) (Figure 4A, Additional File 2 - Tables S3, S7, S11, S19, and S23). However, these results were not statistically significant. There was strong statistical evidence for increasing odds of asymptomatic infection with increasing compound sero- prevalence for two antigens (Etramp5.Ag1 and *Pf*MSP1_19_) (Figure 4B, Additional File 2 - Tables S4, S24), and weaker statistical evidence for increasing odds of asymptomatic infection for HSP40.Ag1 and Rh2.2030 (Additional File 2 - Tables S14, S16). Average compound size in regions with less than 15% PCR prevalence (West Coast, North Bank, Lower River and Central River Regions) was 9.3 (95%CI 7.9 – 10.7) individuals, and in regions with more than 15% PCR prevalence (Upper River Regions South and North), average compound size was slightly larger (16.3 95%CI 13.4 – 19.1 individuals) (Additional File 1 – Figure S1). Across all compounds, the average age range within a household was 54.2 years (95%CI 51.2 – 57.1), with an average minimum age of 2.6 years (95%CI 2.1 – 3.0) and a maximum age of 56.7 years (95%CI 53.8 – 59.8) (Additional File 1 – Figure S1).

**Figure 4.**
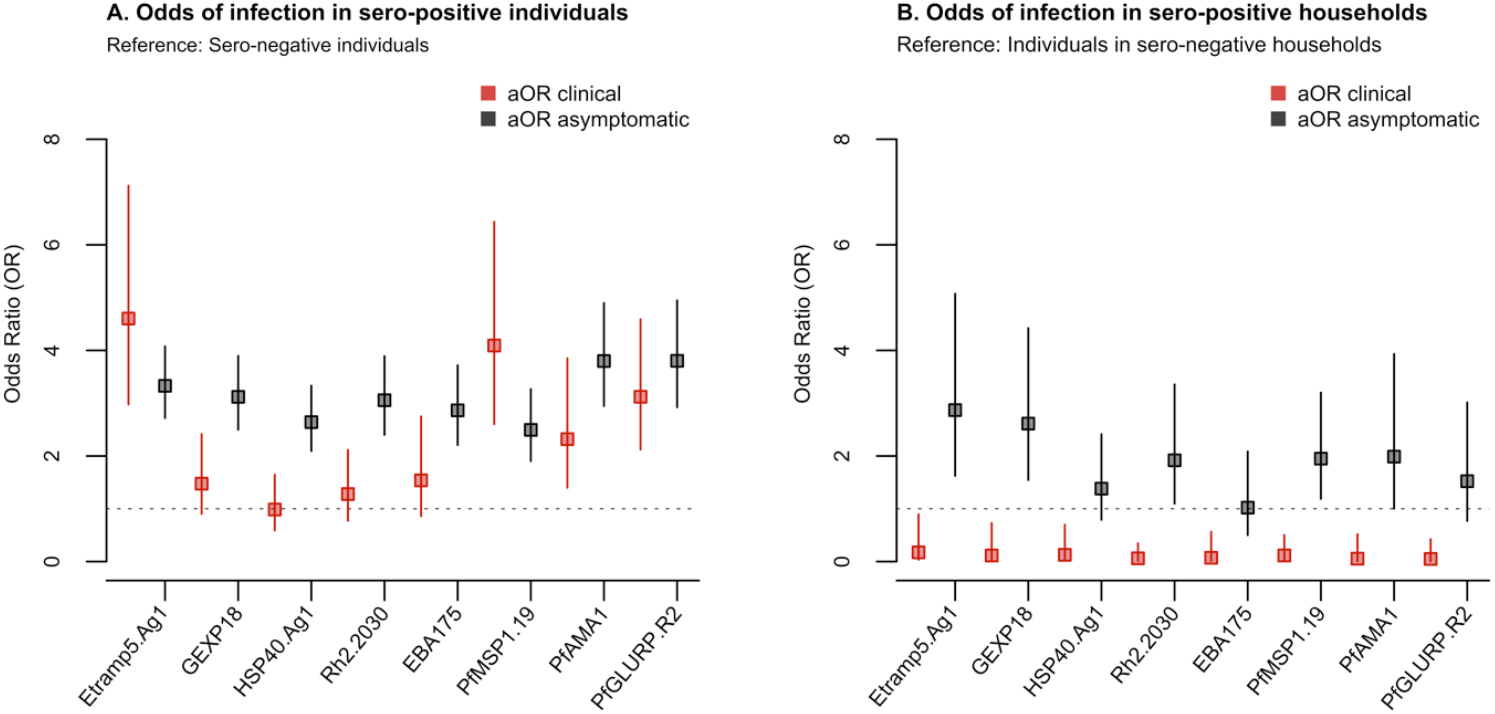
Odds clinical malaria and asymptomatic infection by a) individual sero-positivity status b) residing in the same compound as a sero-positive individual. Analyses is adjusted for age group (<5 years, 5-15 years, and >15 years) and LLIN use in the last 24 hours and weighted by compound size.

Antigens most associated with increased odds of asymptomatic infection were Etramp5.Ag1, GEXP18 and *Pf*MSP1_19_. For Etramp5.Ag1, aOR 2.52 (95%CI 1.49 - 4.28, p=0.001) in compounds with less than 50% sero-prevalence to an aOR 8.17 (95%CI 5.23 - 12.76, p<0.001) in compound sero-prevalence greater than 50%, compared to sero-negative compounds (Additional File 2 - Table S4). For GEXP18, aOR ranged from 2.86 (95%CI 1.44 - 5.69, p=0.003) to 8.85 (95%CI 3.54 - 22.08, p<0.001) (Additional File 3 - Table S9). For *Pf*MSP1_19_, aOR ranged from 2.13 (95%CI 1.30 - 3.50, p=0.003) to aOR 8.75 (95%CI 4.96 - 15.43, p<0.001) (Additional File 2 - Table S24). For HSP40.Ag1, EBA175, and *Pf*AMA1, statistically strong evidence for increased odds of asymptomatic infection was only observed in compounds with greater than 50% sero-prevalence (Additional File 2 - Tables S12, S20, S28).

The spatial distribution of infections at both the start and end of the transmission season (June – July 2013 vs August-December 2013) suggest that clinical and asymptomatic malaria are localised near households with Etramp5.Ag1 sero-positive individuals (Figure 5). There are a number of Etramp5.Ag1 sero-positive households at both the start and during the wet season without concurrent clinical or asymptomatic infections present. However, this is more pronounced for *Pf*AMA1, where sero- positivity could reflect a longer history of exposure than Etramp5.Ag1 (Additional File 3 - Figure S1). While there are a limited number of clinical cases between June and July 2013, from August to December 2013, these cases occur in proximity to households with asymptomatic and Etramp5.Ag1 sero-positive individuals.

**Figure 5.**
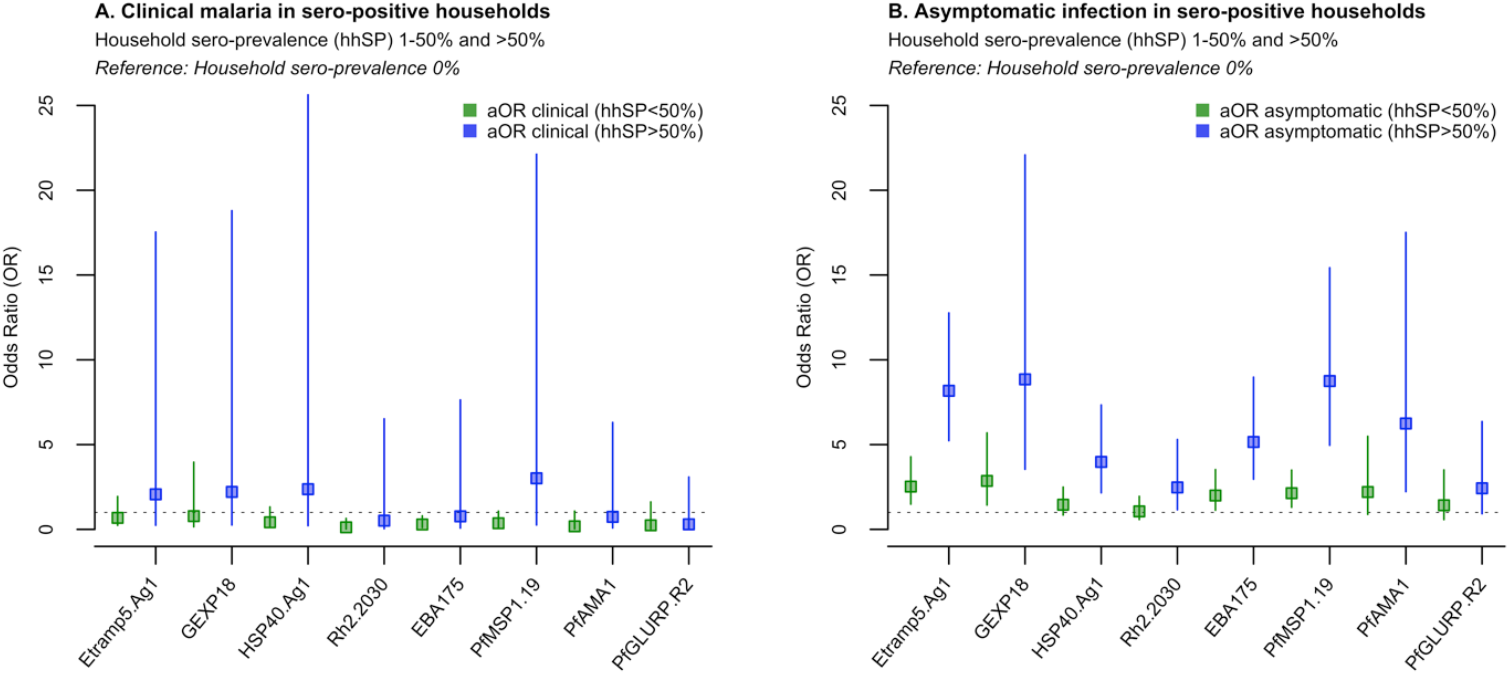
Odds of clinical malaria by compound sero-prevalence (a) and odds of asymptomatic infection by compound sero-prevalence (b). Analyses is adjusted for age group (<5 years, 5-15 years, and >15 years) and LLIN use in the last 24 hours and weighted by compound size.

**Figure 6.**
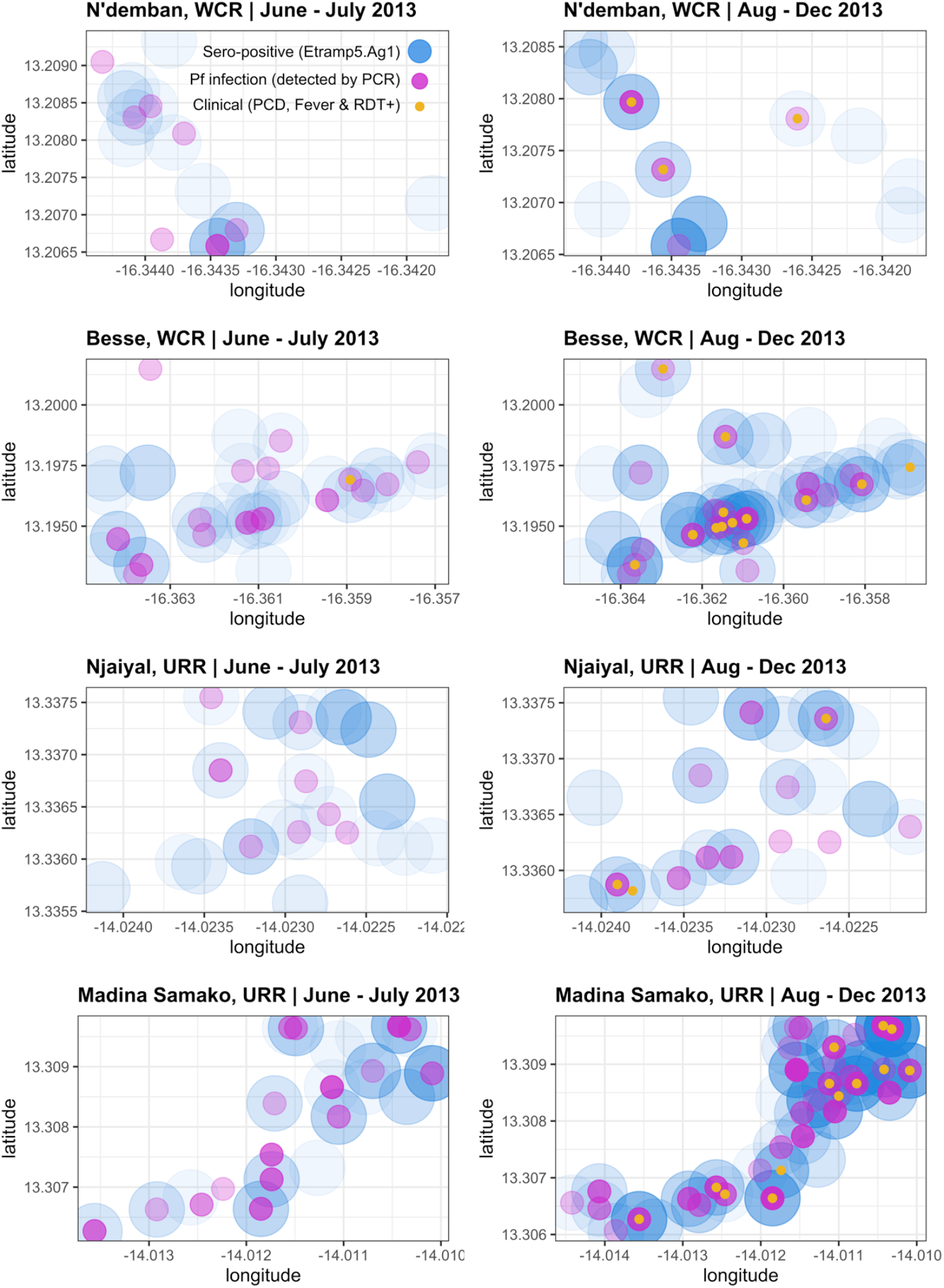
Household geolocation of Etramp5. Ag1 sero-positive, clinical malaria and *Pf* infections across four villages. Spatial distribution of infections shown for lower transmission (N’demban) and higher transmission (Besse) villages in the West Coast Region (WCR) and lower transmission (Njaiyal) and higher transmission (Madina Samako) villages in the Upper River Region (URR). Infections at the start of the wet season (June – July 2013) shown on the left and during the wet and transmission season (August – December 2013) on the right.

## Discussion

We investigated the association between antibody responses and clinical and parasitological endpoints at the individual, compound, and population level in The Gambia. At the individual level, clinical malaria was highly correlated with concurrent sero-positivity to Etramp5.Ag1, *Pf*MSP1_19_, *Pf*AMA1, and *Pf*GLURP.R2, while asymptomatic infection was correlated with sero-positivity to all antigens included in this study. The strongest associations were with sero-positivity to Etramp5.Ag1 and *Pf*AMA1. Village-level sero-prevalence amongst children 2-10 years against antigens Etramp5.Ag1, HSP40.Ag1, and *Pf*MSP1_19_ showed the highest correlations with clinical incidence and *Pf* infection rates.

At the compound level, individuals had increased odds of asymptomatic infection when residing in a compound with greater than 50% sero-prevalence to all antigens, with the highest odds associated with sero-prevalence to Etramp5.Ag1, GEXP18, and *Pf*MSP1_19_. These results do not imply a causal relationship between serological responses and clinical outcomes or asymptomatic infection, but highlight that infected individuals tend to be concurrently sero-positive to most of the serological markers assessed. There was strong evidence of reduced odds of clinical malaria for individuals residing in the same compound as an individual sero-positive to any of the antigens, suggesting a protective effect. In fact, previous studies suggest that several of the antigens investigated in this study – EBA175, Rh2, *Pf*GLURP.R2, *Pf*AMA1 and *Pf*MSP1 – have a functional role in mediating protective immunity if antibodies are acquired at a sufficiently high magnitude.(21–23)

While *Pf*MSP1_19_ and *Pf*AMA1 have been used extensively to measure medium and long-term trends in transmission intensity, these markers may be less sensitive to changes in transmission at higher endemicities or to smaller short-term fluctuations due to the rapid acquisition of antibodies and the long half-life of sero-positivity for some blood-stage antigens.(24–27) Individuals with clinical malaria or with long-lived antibody responses in low transmission regions of western Gambia could indicate foci of malaria transmission that have been ongoing for several transmission seasons or years. In areas where parasite density is low and often missed by RDTs and microscopy, using serological markers could support targeted interventions to clear or eliminate these residual reservoirs of human infection. Antigens in this study associated with shorter-lived sero-positivity (Etramp5.Ag1, GEXP18, and HSP40.Ag1) were positively correlated with either clinical malaria and asymptomatic infection, and sometimes both. These antigens have been relatively recently identified and shown to decrease rapidly after infection in Ugandan children.(28) The cellular localisation of these antigenic targets make them less likely to play a central role in mediating functional immunity, unlike antigens associated with longer-lived antibodies such as *Pf*AMA1, *Pf*MSP1_19_ and EBA175. In settings where most infections are sub-patent and fluctuate above or below the detection limit of PCR, detecting the short-lived antibody response in individuals or households could imply recent infectivity, particularly in the dry season where clinical cases are rare.

While factors such as the method for determining sero-positivity cut-off values could affect estimates of sero-prevalence or the magnitude of association between endpoints,(29) the observations in this study have important implications. In particular, there is a very consistent trend of compound sero- positivity being a risk factor for asymptomatic infection in contrast to being protective against clinical malaria. The complex dynamics between clinical and parasitic immunity are not explained through epidemiological data alone as previous studies have observed lower PCR density in areas of higher transmission intensity(30). Fluctuations in sero-positivity levels may be a strong indicator of a persistent human reservoir of infection, warranting community-targeted interventions if individuals remain infected but are not clinically apparent and continue to transmit. Household serological status could be used for targeting interventions where there are foci of asymptomatically infected individuals.

Immuno-epidemiological studies will be subject to variations in antigen-specific antibody dynamics, as well as the immunogenicity of recombinant antigen constructs. Differences in sequence selection and expression systems may result in recombinant proteins that differ slightly from natural epitope conformation or exposure. It is unclear the degree to which these subtle variations affect the strength of antibody detection between recombinant antigens. While this study focuses primarily on total IgG responses, quantification of antibody levels using a combination of isotypes or subclasses may allow for improved detection of sero-incidence. For example, some studies have found that IgG3 antibodies bind more weakly or have a reduced serum half-life relative to IgG1(31,32), suggesting that naturally acquired antibody responses can be skewed with respect to isotype and subclass. Studies on switch class variants for both malaria and other infectious pathogens have found that antibody affinity and avidity will differ between antigens of the same pathogen or even between different domains of a single antigen(33,34). This can be due to factors such as age and gender, antigen density, strain of the organism, and epitope specificity(32,35,36). Ultimately, current evidence on the associations of antigen-specific IgG subclasses with both exposure and immunity to malaria is varied(37,38). Antigens will fall along a continuum of suitability as either biomarkers of acute infection to correlates of protective immunity(22), and these subtleties need to be considered when selecting targets for surveillance. The assays used in this study have allowed us to quantify antibody responses to specific antigens. Therefore, it will also be valuable to investigate the optimal combinations of antigens associated with both short-lived and long-lived antibody responses in surveillance, which could be more routinely used to account for the breadth of antibody response between individuals in a population.(39–41) Variations such as this could also potentially be overcome by the use of multiple variants or chimeric antigen constructs.(42) Additionally, quantifying the half-life of these antibody responses may help to inform how these antigenic targets can be used to estimate time since last infection.

Changing household sero-prevalence throughout the season may influence the precision of estimates, which could be overcome with more frequent sampling of study participants, such as three-month intervals depending on the decay rate of antigen-specific antibodies. The findings in this study suggest that household sero-prevalence may be a robust proxy for past malaria exposure. It is simultaneously associated with increased risk of asymptomatic infections, but reduced risk of clinical malaria (potentially due to partial immunity). However, the biological relationship between household serological status and the risk of clinical or parasitic infection could be better investigated and confirmed through controlled observational cohort studies.

These findings indicate that a number of new serological markers of malaria exposure could be useful for epidemiological surveillance in highly seasonal malaria transmission areas. There is potential for several of the antigenic targets explored to be used for population-wide screening, all of which have also been optimised for use on a multiplexed bead-based platform in research settings. Further translating this to a lateral flow device may offer opportunities for more widespread application if use-cases can be well established.(43) In light of the strong association between serological status and increased asymptomatic infection, serological platforms may have direct utility as a screening tool in active or reactive case detection strategies, most of which have relied on clinical index cases presenting at health facilities and have shown variability in effectiveness.(44) Future work can also aim to develop standardised methods for characterising serological responses at the village or cluster level or as part of randomised trials, while optimal sampling timeframes and sentinel populations can be designed for use in routine surveillance.

## Conclusions

As more countries move towards malaria elimination, including several in sub-Saharan Africa, sensitive surveillance tools will be needed to further understand the dynamics between the parasite, host, and vector at very low levels of transmission. Integrating newly developed serology markers of recent exposure into existing field diagnostics and case reporting will become more important in estimating the human reservoir of malaria infection, understanding patterns of transmission, and informing the targeting of elimination strategies.

## Data Availability

The datasets used and/or analysed during the current study are available from the corresponding author on reasonable request.

## List of abbreviations

aOR: Adjusted odds ratio
CI: Confidence interval
DBS: Dried blood spot
EBA175: Erythrocyte binding antigen 175
Etramp5.Ag1: Early transcribed membrane protein 5
GEE: Generalised estimating equations
GEXP18: Gametocyte export protein 18 (GEXP18)
HSP40.Ag1: Heat shock protein 40
IgG: Immunoglobulin
LLIN: Long-lasting insecticide treated net
MDA: Mass drug administration (MDA)
MFI: Median fluorescence intensity
PCD: Passive case detection
PCR: Polymerase chain reaction
*Pf, P.falciparum*: *Plasmodium falciparum*
*Pf*AMA1: *P.falciparum* apical membrane antigen 1
*Pf*MSP1_19_: *P.falciparum* merozoite surface antigen 1 19-kDa carboxy-terminal region
RDT: Rapid diagnostic test
Rh2.2030: Reticulocyte binding protein homologue 2
URR: Upper River Region, The Gambia
WCR: West Coast Region, The Gambia

## Declarations

## Acknowledgements

[Not applicable]

## Funding

This study was funded by the UK Medical Research Council (UKMRC) through the LSHTM Doctoral Training Programme studentship received by LW. The funders had no role in the design of the study, collection, analysis, interpretation of data or writing of the manuscript.

## Consent for Publication

[Not applicable]

## Author contributions

LW designed and coordinated the study, supervised and performed serological lab work, analysed the data, and wrote the manuscript. JM and MA supervised field data collection and laboratory work. SC and MB conducted the serological assays. KKAT and JB produced the antigens. KKAT and TH developed the serological assay. JM, MA, UD, IK, and CD advised on study design, interpretation of findings, and reviewed the manuscript. All authors read and approved the final manuscript.

## Competing interests

All authors declare no competing interests.

## Additional files

**Additional File 1. Sero-prevalence amongst individuals aged 2 - 10 year, prevalence of sero-positive compounds, and population size and age distribution by compound**.

**Additional File 2. Odds of clinical malaria and asymptomatic infection for sero-positive individuals and individuals residing in sero-positive compounds**. Unadjusted and adjusted odds ratios for eight malaria antigenic targets - Etramp5.Ag1, GEXP18, HSP40.Ag1, Rh2.2030, EBA175, *Pf*MSP1_19_, *Pf*AMA1, and *Pf*GLURP.R2.

**Additional File 3 – Figure S2. Map of household geolocation of *Pf*AMA1 sero-positive, clinical malaria and *Pf* infections across four villages during dry and wet transmission seasons**. Spatial distribution of infections shown for lower transmission (N’demban) and higher transmission (Besse) villages in the West Coast Region (WCR) and lower transmission (Njaiyal) and higher transmission (Madina Samako) villages in the Upper River Region (URR). Infections at the start of the wet season (June – July 2013) shown on the left and during the wet and transmission season (August – December 2013) on the right.

## Notes

### Competing Interest Statement

The authors have declared no competing interest.

### Funding Statement

This work was supported by the UK Medical Research Council (UKMRC) through the LSHTM Doctoral Training Programme studentship received by LW. The funders had no role in the design of the study, collection, analysis, interpretation of data or writing of the manuscript. 

